# Impact of climate change on human health: Exploring challenges and opportunities through a qualitative study in Birhan Field Site, Amhara Region, Ethiopia

**DOI:** 10.1101/2025.04.26.25326478

**Authors:** Chalachew Bekele, Anum S. Hussaini, Addisalem Fikre, Clara Pons Duran, Yahya Mohammed, Bezawit M. Hungenaw, Bryan Vu, Grace Chan

## Abstract

Ethiopia’s climate exhibits significant regional variability, with varying rainfall patterns and consistently rising temperatures across the country. Climatic changes have direct and indirect effects on human health by increasing natural disasters such as flooding and droughts, increase food insecurity through reduced crop yields, and increase infectious disease transmission. Significant gaps exist in climate-health research, compounded by insufficient funding and a dependence on international evidence for local policy development. We explored community perspectives on the impact of climate variations on the environment, health and livelihood of people in Ethiopia to prioritize and inform mitigation strategies. We conducted a qualitative study in Ethiopia to discuss effects of climate change on human health, livelihoods, and the environment, as well as potential mitigation strategies for the country. Health workers, public health experts, and representatives from various organizations were interviewed. We systematically coded the data to identify prominent and emerging themes. We identified three overarching themes: (I) drivers of climate disruption and environmental degradation, (II) socioeconomic impacts of climate change on communities, (III) strategies for mitigating climate change and its impact. Key findings reveal weather pattern shifts, reduced crop yields, and environmental degradation caused by urbanization and improper waste disposal. Participants shared concerns including weather impacts on crop production and rising health risks, and proposed solutions including afforestation and reducing carbon emissions. Further research is needed to address climate disruptions, especially in low-resource, industrializing areas. Our findings underscore the imperative for collaborative efforts among policymakers, researchers, and local communities to address the multifaceted challenges posed by climate change. Future research should prioritize developing evidence-based policies and interventions tailored to local contexts, aiming to enhance resilience and minimize the adverse impacts on Ethiopia’s population and ecosystems. By leveraging these insights, we can work towards a sustainable future that mitigates climate risks while fostering socio-economic and environmental stability.

## Introduction

Climate change represents one of the most significant challenges facing humanity in the twenty-first century, contributing to an increase in humanitarian emergencies (1). Climate change is characterized by an increase in extreme weather events, including the increased frequency and severity of droughts, heat waves, intense rainfall events, and floods and manifested through increased aridity and water scarcity, dust storms, tropical cyclones (typhoons or hurricanes), wildfires, the melting of snow, and the disappearance of river deltas and coastal cities (2,3). These events are escalating in both scale and intensity, and future climate projections indicate a continued rise in global temperatures (4). These trends are expected to exacerbate the impacts of climate change on natural and human systems, predicting a global increase of approximately 250,000 additional deaths annually between 2030 and 2050 due to undernutrition, malaria, diarrhea, and heat stress (5). Changes in climate have already initiated a broad spectrum of health issues (6), impacting human health both directly and indirectly. Direct effects include mortality and injuries resulting from floods and storms, while indirect effects involve alterations in the distribution of disease vectors such as mosquitoes, the prevalence of water-borne pathogens, water and air quality, and the availability and quality of food (7). These health impacts are particularly pronounced in lower-income populations, predominantly situated in tropical and subtropical regions (6). Additional indirect effects include climate-induced economic dislocation and environmental decline (6) which disrupt agricultural productivity, displaced populations, and degrade natural resources, leading to significant economic and social challenges, contributing to long-term vulnerabilities, particularly in regions with limited adaptive capacity (5).

The continent of Africa is highly susceptible to future climate change, with Ethiopia as one of the extreme examples (8). Ethiopia’s climate varies significantly across different regions. The southeastern and northeastern lowlands experience a tropical climate, while the highland regions have much cooler temperatures (9). A review of long-term climate data for Ethiopia reveals divergent rainfall trends, with some regions experiencing increased rainfall while others face decreases. However, temperatures are rising uniformly across all regions (10). According to a report by the World Bank Group, the temperature in Ethiopia has increased by approximately 0.2°C per decade over the past several decades. The average annual temperature in Ethiopia is 22.6°C, with monthly temperatures ranging from 20.9°C to 23.9°C (11). Additionally, precipitation is expected to decline from an annual average of 2.04 mm/day in 1961-1990 to 1.97 mm/day in 2070-2099, amounting to a cumulative decrease of 25.5 mm/year (12). Some regions of the country experience frequent droughts that lead to a shortage of grazing areas, grasslands, and water. This scarcity adversely affects food production, resulting in diminished nutritional status and fertility among lactating animals, and a decline in overall health and conception rates, leading to lower household-level production of milk and milk by-products (13). These climatic changes are anticipated to exacerbate food insecurity, leading to malnutrition, cause land degradation, and inflict damage on infrastructure (12,14–17) and spur outbreaks of vector-borne diseases such as malaria, dengue fever, Cholera, and Dysentery, lengthening their transmission periods (14,18). Malnutrition, food insecurity, and child growth and developmental delays are among the major impacts of climatic influences on human health in northeastern Ethiopia (18).

In global health, climate alterations significantly impact challenges like infectious disease control. Current global health efforts prioritize infectious diseases and maternal and child health, often overlooking climate impacts (19). A review conducted in 2016 highlighted that research on climate change and health in Ethiopia is inadequately developed, with few spatially detailed and methodologically consistent studies. This lack of comprehensive local data has led to policies and strategies that are not sufficiently grounded in Ethiopia-specific evidence, often relying instead on international findings (13). There is a shortage of trained researchers and competing priorities has left Ethiopian health policies under informed and reactive rather than proactive (13). Our study seeks to build the local evidence base necessary for sustainable, context-specific public health responses.

The proposed study seeks to describe the impact of climate change among community members in Ethiopia. The objective of this study is to describe current climate impacts, identify gaps and needs related to the impact of climate change on human health and livelihoods, and highlight opportunities to enhance climate change research, training, and capacity-building initiatives in Ethiopia, drawing on the experiences and perspectives of community members.

## Method

### Study design and setting

We conducted a qualitative study at the Birhan Health and Demographic Surveillance System (HDSS) to understand the community perceptions of climate change and its effect on health and environment in Ethiopia. The HDSS is located in the North Shewa zone of Ethiopia, approximately 130 km north of the capital city, Addis Ababa (20). The catchment area includes two districts, Angolela Tera and Kewet/Shewa Robit, encompassing a population of 79,653 individuals and 19,957 households. Each district is divided into eight clusters (kebeles).

The Birhan HDSS is particularly important for climate-health research due to its ecological and demographic diversity. It encompasses both highland and lowland terrains, with altitudes ranging from 1,093 to 3,030 meters above sea level, leading to significant variations in temperature and rainfall patterns. This diversity makes it representative of broader environmental and socioeconomic conditions in Ethiopia. The population is predominantly rural, with livelihoods primarily based on rain-fed agriculture and semi-pastoralism, sectors highly sensitive to climate variability. Additionally, the Birhan HDSS provides comprehensive, high-quality longitudinal data on health and demographic indicators, including maternal and child health, water and sanitation access, and disease prevalence. This robust data infrastructure enables in-depth analysis of how climate change impacts health outcomes in vulnerable communities, thereby informing targeted interventions and policy decisions.

### Study population

Using a purposive sampling approach, local healthcare providers, in public health facilities, and various organizations’ representatives working in the area were invited to discuss their knowledge and awareness of climate change and its impacts on study population life. The discussion centered on concerns about climate change, particularly its impact on human health and livelihoods. Participants reviewed current efforts to address these challenges within the community and shared suggestions for future strategies to mitigate and adapt to climate change in their region.

### Data collection

We conducted two Focus Group Discussions (FGDs) in January 2023. Participants were interviewed using semi-structured guides tailored to inform the selection of interview topics. The FGD questions were designed to capture four broad categories: participants’ knowledge and perceptions of climate change, factors responsible for climatic fluctuations, its effects on the environment, health and livelihood, and potential mitigation strategies to minimize these impacts of climate change.

Each FGD was conducted with all participants present in the same room to promote shared dialogue and ensure consistency in the discussion. All participants were asked the same set of questions, and care was taken to provide each individual with an opportunity to express their views and experiences. The duration of FGDs ranged between 40 to 60 minutes. The FGDs were facilitated in Amharic and audio recorded for transcription purposes, with permission from respondents. Field notes were recorded during all interviews to capture contextual details and facilitate comprehensive analysis. Interviews were transcribed in English and the quality of all transcripts were closely monitored. The clarity and accuracy of participants’ responses were ensured through summarization and paraphrasing techniques.

### Data Analysis

Qualitative transcripts were analyzed using a thematic analysis approach. This approach offers considerable flexibility in both theoretical conceptualization and methodological design, allowing for a comprehensive examination of patterns and themes within the data, accommodating various research frameworks and objectives (21). The ATLAS.ti software (version 23) was used to import, organize, explore, and code data for analysis. First, deductive codes derived from the interview guide were systematically applied to the transcripts to identify prominent themes. Subsequently, inductive codes were added to the transcripts to capture emerging themes. These themes were derived from the four major categories under which the questions were arranged in the semi-structured FGD guide. This systematic approach ensured consistency and comprehensiveness in coding. All queries concerning the transcripts’ textual content were discussed with the research team in the consensus meetings to refine interpretations and coding decisions and reduce researcher biases. All preliminary themes were subsequently grouped according to the research question. Following that, memos were written, describing coding categories, emergent themes, and relationships between constructs and codes.

## Results

Forty-nine participants were interviewed for the study. The participants included both males (n=36) and females (n=13). The sample encompassed a variety of professional roles, including department heads (n=7), facility/health center heads (n=12), health extension supervising officers (n=2), Woreda/district leads (n=2), health supervisors (n=9), Kebele/village managers (n=11), and religious leaders (n=6). Participants exhibited a range of sociodemographic characteristics, which provided a comprehensive overview of the diversified perspectives within the community.

Through thematic analysis, we identified three overarching themes (Figure 1): (I) drivers of climate disruption and environmental degradation, (II) socioeconomic impacts of climate change on communities, and (III) strategies for mitigating climate change and its impact. Figure 1 summarizes the major findings by themes with supporting quotes from the FGDs.

**Figure 1:**
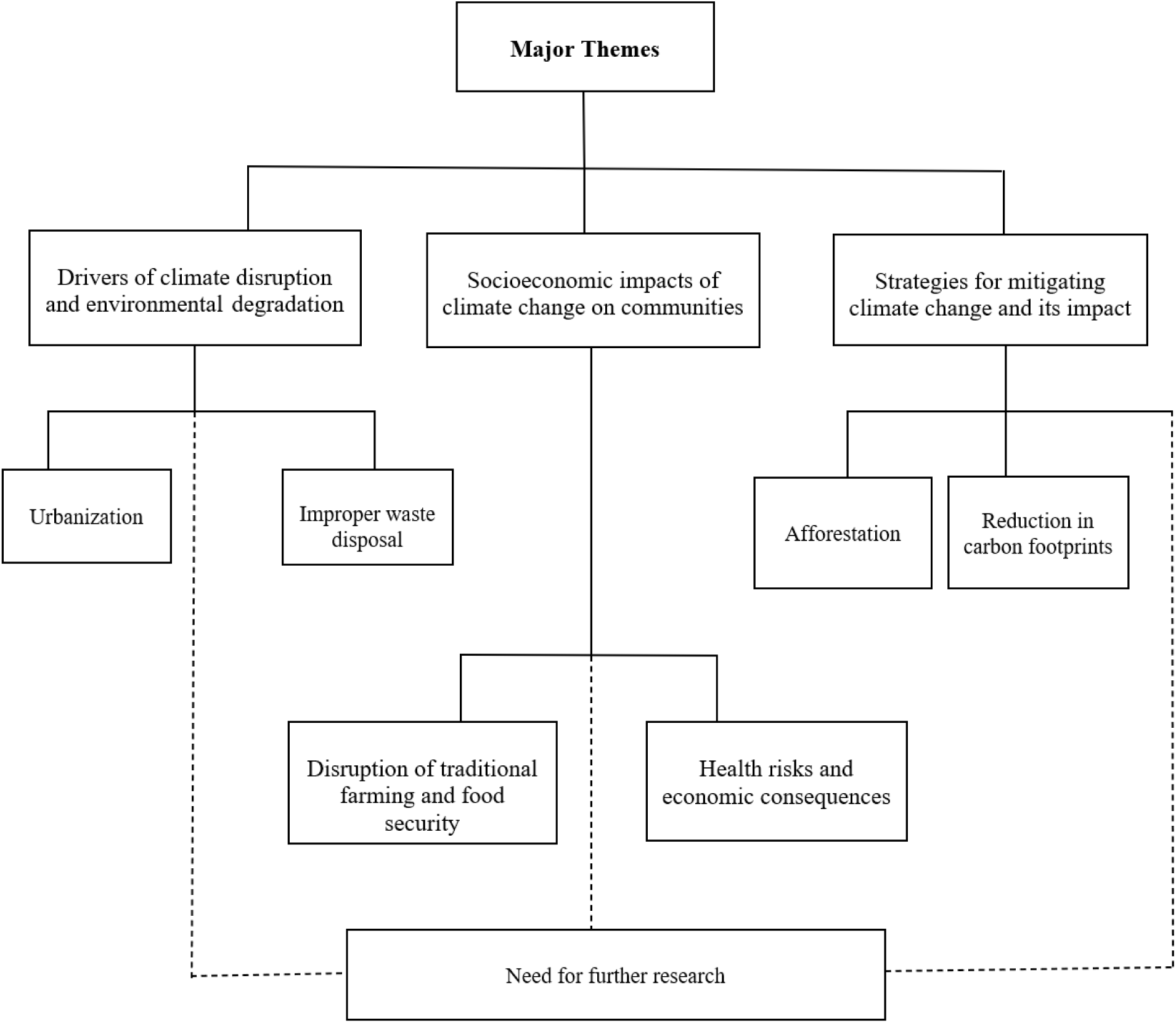
Flowchart of major themes extracted from the FGDs

### **(I)** Drivers of climate disruption and environmental degradation

#### (a) Urbanization

Participants emphasized urbanization as a primary driver of climate variability. The expanding population has intensified environmental pollution and deforestation for housing and the expansion of industries, factories, retail, and healthcare services. Increased technological use, particularly vehicles, has further amplified carbon footprints, contributing to climate fluctuations. Approximately 85% of the vehicle fleet in the country comprises imported second-hand vehicles, many of which are diesel-powered and over 20 years old. These aging vehicles often lack modern emission control technologies, leading to high levels of pollutants such as particulate matter (PM), nitrogen oxides (NOx), and carbon monoxide (CO).

“*The city of Debre Birhan was once very cold, but the temperature has changed due to the increase in population and the presence of more factories in the area.*”

“*In the past, cold weather in our area (Debre Birhan) would often break water meters, but now the weather is becoming more similar to that of Shewa Robit, which is known for being hot. This change in weather may be due to industrial expansion in the area.*”

#### (b) Improper waste disposal

Participants emphasized that improper waste disposal from factories and towns often ends up in rural areas. This issue was linked to the degradation of local waterways, with several participants noting the accumulation of plastic and industrial waste in rivers and primary water sources flowing through the Birhan being impacted by effluents from nearby industrial activities, including tanneries in Debre Birhan. The wastewater discharged into the local water sources contains high concentrations of pollutants and other heavy metals, contributing to environmental degradation and posing risks to downstream communities. Participants reported that such pollution reduces water quality, decreases land productivity, and contributes to the rise in waterborne and vector-borne diseases. Livestock and cattle, which are vital to household livelihoods in the area, were also noted to suffer adverse effects from consuming contaminated water or grazing on affected land. Inadequate waste management systems and limited environmental monitoring were identified as contributing factors, amplifying the negative impact on human health, agricultural sustainability, and overall ecological resilience in the region.

“*There is no monitoring or regulation to ensure that waste is properly disposed of. As a result, the community in Chacha and Chefanen kebeles have reported that the improper disposal of waste is killing their cattle.*”

#### (c) Need for further research

Participants discussed human practices impacting the natural environment, focusing on industrial expansion’s consequences and its ripple effect on community livelihoods, particularly cattle well-being. They proposed a comprehensive study to understand and address these challenges, aiming to develop informed plans that mitigate industrial expansion’s adverse effects on cattle, demonstrating a commitment to resolving the intersection of human practices and environmental impact.

“*In the past, cold weather in our area (Debre Birhan) would often break water meters, but now the weather is becoming more similar to that of Shewa Robit, which is known for being hot. This change in weather may be due to industrial expansion in the area.*”

### **(II)** Socioeconomic impacts of climate change on communities

#### (a) Disruption of traditional farming and food security

Participants noted significant impacts from recent weather changes on traditional farming reliant on seasonal rainfall for crop production. They observed unpredictable weather patterns, including unexpected temperature shifts in historically warm regions, affecting crop types grown. Climate fluctuations have detrimentally affected crop yields, with heavy rainfall destroying crops and diminishing the effectiveness of farming inputs like urea. Additionally, depletion of water sources have raised concerns over clean water shortages for drinking, hygiene, and irrigation, forcing community members to purchase water at high prices. One participant reported paying 25 Ethiopian Birr (ETB) per jerrican of water. Given that a standard jerrican holds 20 liters, this equates to 1.25 ETB per liter, an amount that is up to 700 times higher than municipal rates in urban centers like Addis Ababa, where water costs between 0.00175 and 0.02671 ETB per liter. This significant disparity exemplifies how climate-induced water scarcity is devastating household economies in rural regions, redirecting resources away from food, healthcare, and education. These compounding pressures have led to reduced harvests, exacerbated inflation, diminished purchasing power in both rural and urban markets, and strained family livelihoods—ultimately deepening the socioeconomic impacts of climate change.One participant reported, “*Climate change is a serious concern. In the past, areas with temperatures below 80 were called “kola” and produced crops like sorghum, beans, and barley. However, now the area called “bukait” is not producing these crops, which is likely due to climate change.*”

Another stated, “*I recently visited Rasa (Kewet Woreda) and spoke with a father who told me that, in the past, there was no shortage of drinking water in the area. The seasonal rivers stayed full for a long time and there were no issues with crop production due to prolonged rainy seasons. However, he told me that the weather has changed significantly in recent years. The rainy season is now shorter, making it difficult to access drinking water and forcing residents to purchase water for 25 ETB per jerrican.*”

#### (b) Health risks and economic consequences

Participants discussed the health impacts of climate change, emphasizing water contamination as a route for infections and changes in disease patterns. They highlighted risks like respiratory infections, waterborne diseases, heat-related illnesses, and sun exposure-related cancers. Economic consequences such as displacement were noted to affect psychological health, while food insecurity and malnutrition posed risks, especially among vulnerable groups. Environmental factors like ozone depletion were cited for exacerbating health issues, including increased mortality due to climate-induced illnesses.

One participant stated, “*In the past, we could expect sunny and warm weather during the harvest season, but now we are experiencing windy, rainy, and cold conditions. This change in weather also had an impact on our health, as many children and adults are experiencing flu-like symptoms and sinus-related diseases. It is clear that the weather has changed significantly in recent times*.” In Ethiopia, the primary harvest season, known as the Meher season, typically occurs from September to November, following the main rainy period from June to August. This season is crucial for food security, as it accounts for the majority of the country’s annual crop production.

Another mentioned, “*In terms of health impacts, previously uncommon diseases like malaria may become more prevalent in highland areas due to rising temperatures that transform these regions into lowlands.*”

#### (c) Need for further research

Participants noted farmers’ frequent use of pesticides and insecticides to enhance production but expressed concern about farmers’ unawareness of the health risks associated with these chemicals. They emphasized the necessity for further research to assess these risks and develop educational programs. Additionally, they stressed the need for comprehensive, evidence-based research on climate change’s impact on agriculture, citing increased cancer rates linked to climate factors. Participants underscored bridging the gap between agricultural practices and scientific knowledge to safeguard farmer and consumer well-being. They advocated for informed decision-making in promoting sustainable and health-conscious farming practices.

“*Scientific studies conducted globally show that global warming and climate change are causing increasing harm to populations. It would be beneficial if similar studies were conducted in our country, particularly in areas […] where there are industries and in areas where crop production is common. Overall, it is important to bring attention to the impacts of climate change in our region and work towards solutions*.”

### **(III)** Strategies for mitigating climate change and its impact

#### (a) Afforestation

Participants supported afforestation as a proactive solution to climate disruption. They highlighted planting trees strategically in deserted areas to mitigate environmental risks, such as flooding, and enhance resilience to natural disasters. Afforestation was also viewed as an effective way to combat climate change by utilizing forests’ capacity to absorb carbon dioxide to reduce atmospheric carbon levels and help mitigate global warming.

“*In areas with forests, there is cool air and clouds. These areas also have a variety of birds. If we plant trees in desert areas, the weather will change.*”

***(a)*** Reduction in carbon dioxide emission

Participants identified fossil fuel combustion for electricity, heat, and transportation as the primary source of carbon dioxide emissions. They recommended adopting energy-saving strategies to reduce carbon footprints, such as using fuel-efficient vehicles, improving building insulation, and deploying energy-efficient electrical appliances.

“*We can take steps to reduce carbon dioxide emissions, such as using materials or transportation that have a lower carbon footprint or avoiding them altogether.*”

#### (c) Need for further research

Participants highlighted climate change’s disproportionate impact on low-resource settings, noting inadequate efforts to address these challenges. They suggested that understanding these implications is crucial for effective climate crisis management and emphasized the necessity for research, especially in industrial regions, given industries’ significant role in environmental dynamics.

“*Climate change is a global concern that affects countries around the world, including ours. It particularly impacts third world and sub-Saharan countries, both economically and in terms of health impacts. Despite the significant impacts of climate change, little has been done to address it. In our country, we see that leaders and policy makers are debating the issue, but there is a lack of evidence-based research on the impacts in our region.*”

## Discussion

This study identifies the causes of climate fluctuations, their impacts, and effective mitigation strategies. Key findings highlight the extensive effects of climate change on population health, including water shortages from reduced rainfall, leading to costly clean water purchases. For instance, residents in affected areas report paying 25 Ethiopian Birr (ETB) per 20-liter jerrican of water. Considering that the average monthly income for rural households in Ethiopia is approximately 7,985 ETB (22), this expense represents a significant financial burden, especially for families requiring multiple jerricans daily. Such costs can consume a substantial portion of household income, diverting funds from other essential needs like food, education, and healthcare. The study also notes the health risks of improper waste disposal and inefficient waste management. Afforestation and energy-saving methods were recommended to combat climate disruption and reduce fossil fuel carbon emissions. These strategies aim to decrease energy consumption and lower carbon dioxide levels, promoting sustainability. Additionally, the study underscores the disproportionate impact of climate change on low-resource areas, particularly in industrial regions, where inadequate infrastructure and inefficient resource management exacerbate vulnerability. This highlights the need for focused research to develop targeted mitigation strategies for these communities, which often lack the capacity to adapt to climate disruptions.

One study reveals regional variations in Ethiopia’s rainfall: increased in the west, but declining in the central rift valley and the eastern and western escarpments, indicating a need for region-specific strategies (23). Another study reports that since 2006, various Ethiopian regions have faced outbreaks of watery diarrhea, underscoring the importance of clean water access and targeted interventions (13). In Addis Ababa, poor waste management from domestic and industrial activities degrades water quality, highlighting the need for effective waste control (24). In the Birhan area, the Beressa River serves as a vital water source for both domestic and agricultural use. This perennial river originates from the Qundi highlands, approximately 30 km east of Debre Birhan, and flows northwest to join the Jema River, a tributary of the Blue Nile. The river’s catchment area spans 211 km², with discharge rates varying seasonally, peaking at 58.78 m³/s in August and dropping to 0.40 m³/s in January (25). However, rapid urbanization has led to increased pollution levels in the Beressa River, with industrial and domestic waste contributing to its contamination (25). This degradation not only affects water quality but also poses significant health risks to the local population, emphasizing the need for improved waste management and water treatment infrastructure. One study suggests that addressing climate change requires multiple strategies: conventional mitigation, negative emissions technologies, and radiative forcing geoengineering. It also identifies gaps in climate research, noting significant attention on policy and renewable energy but a lack of focus on specific mitigation technologies and geoengineering (26). A recent review also emphasizes the need for innovative, context-specific adaptation strategies to protect vulnerable communities from the adverse health effects of climate change (27).

The study’s findings aim to highlight the need for short and long-term staff training in climate change and health sciences, such as carbon trade negotiation, geospatial analysis, and hazard mapping. It emphasizes the importance of collaborative efforts in research, training, and policy support across health, environment, sociology, and economics to improve risk assessment and management. The findings are crucial to prioritize initiatives like afforestation, community awareness on climate-related health risks, and developing climate change and health curricula for undergraduate and postgraduate programs, and health workers in Ethiopia.

The research has some notable limitations, including a small sample size of two FGDs, which has limited data and robust conclusions. Purposive sampling may have also introduced selection bias and limited result generalizability due to lack of population diversity. The study benefited from knowledgeable public health leaders with extensive professional profiles, providing diverse perspectives. This approach enabled a detailed examination of specific climate change impacts, yielding valuable and practical recommendations for their communities.

## Conclusion

This study underscores the profound impacts of climate change on Ethiopia’s environment and public health, as articulated by local community perspectives, calling for immediate actions and continued research to mitigate the adverse effects of climatic disruptions. Urbanization and improper waste disposal have emerged as significant contributors to environmental degradation, exacerbating challenges such as reduced crop productivity, water scarcity, and increased health risks. The study advocates comprehensive, region-specific research to better understand these dynamics and develop effective adaptation strategies. Afforestation and carbon emission reductions were identified as promising solutions to combat climate destabilization, emphasizing the importance of sustainable practices and informed policymaking. Addressing these challenges requires collaborative efforts across disciplines and sectors to safeguard livelihoods and ecosystems, ensuring a resilient future for Ethiopia amidst ongoing climate crises.

## Data Availability

The datasets used and analyzed during the current study are available at the HaSET on a reasonable request.

## Acknowledgements

We are highly grateful to all the participants for sharing their valuable insights and all the research staff for their support in data collection and entry. We appreciate the valuable insights from the Ministry of Health regarding future research directions and policy priorities.

## Declaration

### Ethics approval and consent to participate

Verbal informed consent was obtained from all participants involved in the study.

## Authors’ contributions

CS, ASH, CPD, and GJC conceptualized and developed the research question. AFT and CS collected the data. ASH conducted data analysis. All co-authors contributed to the writing of the manuscript and provided feedback and approved the final version of the manuscript.

## References

1. World Health Organization. Climate Change 2023 [Internet]. Available from: https://www.who.int/news-room/fact-sheets/detail/climate-change-and-health

2. Handmer J, Honda Y, Kundzewicz ZW, Arnell N, Benito G, Hatfield J, et al. Changes in impacts of climate extremes: human systems and ecosystems. In: Managing the Risks of Extreme Events and Disasters to Advance Climate Change Adaptation. Cambridge (UK): Cambridge University Press; 2012. p. 231–90.

3. Nava A, Shimabukuro JS, Chmura AA, Luz SLB. The impact of global environmental changes on infectious disease emergence with a focus on risks for Brazil. ILAR J. 2017;58(3):393–400.

4. Meehl GA, Stocker TF, Collins WD, Friedlingstein P, Gaye AT, Gregory JM, et al. Global climate projections. In: Climate Change 2007: The Physical Science Basis. Cambridge (UK): Cambridge University Press; 2007.

5. World Health Organization. Health and climate change 2018 [Internet]. Available from: https://www.who.int/news-room/facts-in-pictures/detail/health-and-climate-change

6. Confalonieri U, Menne B, Akhtar R, Ebi KL, Hauengue M, Kovats RS, et al. Human health. In: Climate Change 2007: Impacts, Adaptation and Vulnerability. Cambridge (UK): Cambridge University Press; 2007.

7. Intergovernmental Panel on Climate Change (IPCC). Synthesis Report, Third Assessment Report. 2011.

8. Conway D, Schipper ELF. Adaptation to climate change in Africa: challenges and opportunities identified from Ethiopia. Glob Environ Change. 2011;21(1):227–37.

9. Bezu A. Analyzing impacts of climate variability and changes in Ethiopia: a review. American Journal of Modern Energy. 2020;3:65–76.

10. Energy Group of ECSNCC Network. Renewable energy and climate change nexus in Ethiopia. 2011.

11. The World Bank Group. Climate Risk Profile: Ethiopia. 2020.

12. Kidanu A, Rovin K, Hardee K. Linking population, fertility and family planning with adaptation to climate change: views from Ethiopia. 2009.

13. Simane B, Beyene H, Deressa W, Kumie A, Berhane K, Samet J. Review of climate change and health in Ethiopia: status and gap analysis. Ethiop J Health Dev. 2016;30(1):28–41.

14. Adem A, Bewket W. A climate change country assessment report for Ethiopia. 2011.

15. Adem A, Guta A. Engendering climate change policy and practice in Ethiopia. Addis Ababa: Forum for Environment; 2011.

16. Oates N, Conway D, Calow R. The ‘mainstreaming’ approach to climate change adaptation: insights from Ethiopia’s water sector. Overseas Development Institute Background Note. 2011.

17. Environmental Protection Agency. National report of Ethiopia, the United Nations Conference on Sustainable Development (Rio+20). Addis Ababa: Federal Democratic Republic of Ethiopia; 2012.

18. Afar National Regional State. Programme of plan on adaptation to climate change. 2010 Oct.

19. Louis MES, Hess JJ. Climate change: impacts on and implications for global health. Am J Prev Med. 2008;35(5):527–38.

20. Bekele D, Hailemariam B, Bekele C, Van Wickle K, Tadesse F, Goddard FGB, et al. Cohort profile: the Birhan Health and Demographic Surveillance System. Int J Epidemiol. 2022;51(2).

21. Braun V, Clarke V. Conceptual and design thinking for thematic analysis. Qual Psychol. 2022;9(1):3.

22. Seyoum A, Hundie SK, Delajara M, Anker R. Living income report (with living wage annex) for rural Sidama, Ethiopia [Internet]. Global Living Wage Coalition; 2024 May 1. Available from: https://www.globallivingwage.org/living-wage-benchmarks/living-income-report-for-rural-sidama-ethiopia/

23. Gemeda DO, Korecha D, Garedew W. Evidences of climate change presences in the wettest parts of southwest Ethiopia. Heliyon. 2021;7(9):e08009. doi:10.1016/j.heliyon.2021.e08009

24. Alemayehu T. The impact of uncontrolled waste disposal on surface water quality in Addis Ababa, Ethiopia. Water Resour Dev. 2001;17(4):539–56.

25. Kebeb E. Water resource management and pollution in the Birhan area: case study of the Beressa River. Ethiop J Environ Stud Manag. 2016;9(3):102–14.

26. Haines A, Ebi K. The imperative for climate action to protect health. N Engl J Med. 2019;380(3):263–73.

27. Fawzy S, Osman AI, Doran J, Rooney DW. Strategies for mitigation of climate change: a review. Environ Chem Lett. 2020;18(6):2067–94.

